# Effect of Insomnia on Repetitive Transcranial Magnetic Stimulation Treatment Outcomes for Depression

**DOI:** 10.1101/2023.12.06.23299444

**Authors:** Jamie Kweon, Andrew Fukuda, Polly Gobin, Lamaan Haq, Joshua C. Brown, Linda L. Carpenter

## Abstract

While repetitive transcranial magnetic stimulation (rTMS) is safe and effective for 50-60% of those treatment-resistant depression, it is critical to identify factors to optimize therapy to help those who do not respond. Baseline sleep characteristics have been investigated as a potential predictor of TMS efficacy but results from various studies have been conflicting. We aimed to explore whether baseline sleep quality, specifically insomnia related symptoms, is associated with TMS outcomes in a naturalistic sample of 975 patients receiving a standard course of rTMS from two sites. One site recorded information on concurrent medication use. Among these 353 patients, we also examined whether pharmacological treatment of insomnia affected TMS treatment response. Depression was measured using the 30-item Inventory of Depressive Symptomology Self Report (IDS-SR) in site one and an abbreviated 16-item Quick Inventory of Depressive Symptomology (QIDS) in site two. Sleep disturbances were measured using three sleep-related questions overlapping between the two questionnaires. We found that sleep quality improves after TMS and correlates with improvement in depression. Upon dichotomous categorization of the sample by insomnia and hypnotics use, we found that among those who had significant insomnia at baseline, those not using sleep medications had significantly worse post-treatment IDS-SR scores compared to those receiving pharmacological treatments for sleep (*p*=.021). Together, our results suggest that while baseline insomnia is not associated with response to TMS treatment, treating insomnia may affect the trajectory of TMS therapy. Future prospective studies are necessary to examine the effect of insomnia treatment alongside TMS for depression.

## Introduction

Though treatments for major depressive disorder (MDD) have significantly improved over the years, we have yet to understand why certain individuals improve while others do not. For those who fail to improve with medications – estimated to be potentially up to 30% of patients (Gaynes et al., 2008), transcranial magnetic stimulation (TMS) has emerged as a revolutionary therapy. However, even TMS response rates are limited to 45-60% with remission rates around 30% (Carpenter et al., 2012; Dunner et al., 2014; George et al., 2013). Understanding who will potentially improve with TMS treatment may guide patient selection and provide insights into pharmacologic augmentation strategies.

Repetitive transcranial magnetic stimulation (rTMS) is an FDA cleared treatment for treatment resistant depression. During rTMS, a coil rests on the scalp and current is rapidly discharged through wire coils to generate a focused magnetic wave (Edinoff et al., 2022). Clinical 10-Hz rTMS to treat depression delivers 10 pulses per second to the dorsolateral prefrontal cortex (dlPFC) and is believed to have excitatory modulatory effects (Ziemann, 2004). At the neuronal level, rTMS is theorized to improve depression symptoms through synaptic plasticity (Brown et al., 2021), with evidence in animals and humans that excitatory stimulation recruits key receptors involved in long-term potentiation (LTP).

Sleep disturbances are a well-established symptom of depression that have been correlated with overall depression severity (Sunderajan et al., 2010). Sleep quality has been previously studied as a promising predictor of MDD treatment outcome across modalities, with research finding objective and subjective measures of sleep disturbance to be associated with poor depression treatment response (Andreescu et al., 2008; Dew et al., 1997; Troxel et al., 2012). Patients with depression have demonstrated abnormal REM sleep (Fang et al., 2019) and disrupted sleep architecture (Murphy & Peterson, 2015). Furthermore, studies have demonstrated that non-depressed individuals with insomnia are three times more likely to develop depression than those with healthy sleep (Lowe et al., 2013). Patients with comorbid insomnia and depression also tend to experience longer durations of treatment and lower remission rates across therapies (Fang et al., 2019), suggesting that poor sleep quality may affect the trajectory of treatment for depression.

Not only does insomnia potentially blunt improvement from depression therapies, but alleviating insomnia through targeted treatment has led to improvements in depression in patients with comorbid conditions. In a study comparing the effects of adjunctive cognitive behavioral therapy for insomnia (CBT-I) with antidepressant treatment compared to antidepressants alone on depression symptoms and polysomnography, only those with CBT-I improved on objective sleep measures while the group without CBT-I worsened (Carney et al., 2017). A meta-analysis of 23 studies also suggests a positive effect of insomnia treatment on depression outcomes, though interventions/ populations were highly variable (Gebara et al., 2018). Interestingly, sleep quality has also been found to facilitate plasticity processes (Centorino et al., 2020), suggesting a potential mechanistic interaction between sleep and rTMS treatments. Given such findings, it seems plausible that sleep quality may modulate the trajectory of rTMS treatment for depression.

Despite the breadth of evidence that suggests insomnia influences depression treatment, current literature conveys mixed conclusions on the influence of sleep quality on rTMS response. Lowe et al. (2013) found no relation between baseline insomnia or hypersomnia and rTMS treatment outcome in an analysis of data pooled from four clinical trials using rTMS treatment for depression (Lowe et al., 2013). Brakemeier et al. (2007) reported that patients with worse baseline sleep had greater likelihood of TMS-related improvement; however, they were not able to replicate these findings in a follow-up study (Brakemeier et al., 2007; Brakemeier et al., 2008). One study did find that early insomnia was related to worse outcome, but the result did not survive after adjusting for trial location heterogeneity (Fregni et al., 2006). Together, these suggest baseline insomnia may not be a strong predictor of TMS treatment response. It is important to note, however, that Lowe et al. (2013) combined data from four trials all using varied frequencies, intensities, and number of sessions. These differing TMS parameters may enact disparate effects on brain networks (Caulfield & Brown, 2022), including ones affecting sleep. Although other studies did control for location in analyses, data was pooled from 6 clinical trials that gave only 10 days of treatment (Fregni et al., 2006), in comparison to the average 30-36 days in a standard rTMS treatment course. Finally, no study examined the potential role of sleep modulators, such as sleep mediations, and how this may have impacted rTMS outcome.

To address these gaps, we used naturalistic data with the largest sample size to date to parse out the role of insomnia in rTMS treatment response, controlling for rTMS parameters as well as investigating the role of sleep medications. We predicted that self-report baseline insomnia is not associated with clinical outcome per previous studies (Brakemeier et al., 2008; Fregni et al., 2006; Lowe et al., 2013), and pharmacologic treatment of insomnia modulates depression symptoms during a TMS treatment course.

## Methods

Data were retroactively analyzed from the medical records of 353 naturalistically treated adult outpatients in the Butler Hospital TMS Clinic and 622 patients in the McLean Hospital TMS Clinic receiving their first course of TMS. As a part of Butler Hospital clinic’s standard assessment battery, patients filled out the Inventory of Depressive Symptomatology Self Report (IDS-SR) and Patient Health Questionnaire-9 (PHQ-9). The McLean site collected the Quick Inventory of Depressive Symptomology (QIDS) and PHQ-9. Both sites collected data at interim time points throughout treatment (before and after course and every 5 treatments at Butler, and every 10 treatments at McLean). All patients had a primary diagnosis of moderate-severe MDD without psychotic features and had inadequate or intolerable response to psychotherapy at least two (and in most cases at least four) antidepressant and/or augmentation medications. Patients were evaluated with a psychiatrist specializing in mood disorders. Patients were on stable medication regimen before starting TMS and were instructed to keep regimens stable throughout the course of TMS.

### TMS Protocol

On the first day of treatment, patients underwent a motor threshold procedure to determine the left hemisphere motor hotspot and minimum stimulator intensity required to produce a right-hand finger twitch for >50% trials. At Butler, patients then began a standard 10-Hz treatment protocol delivering 3,000 pulses a day for 6 weeks, 5 times a week, followed by 6 sessions over 3 weeks. Butler patients were treated with a NeuroStar figure-8 coil (Neuronetics, 2003) over the left dorsolateral prefrontal cortex (dlPFC) at a stimulation intensity 120% of their resting motor threshold (MT). In ∼60% of cases, where patients had difficulty tolerating the 10-Hz protocol at 120% MT, they received 5-Hz stimulation (3,000 pulses) on the same location. A minority of patients were transitioned to 1-Hz stimulation over the right dlPFC at some point during their treatment course; only 12 patients received 1-Hz for more than 50% of treatment sessions. For a minority of non-responding patients, the total number of pulses per session was increased to 4000.

McLean TMS patients were treated on one of two device types; A MagVenture B70 figure-8 coil for 5% of patients, and a BrainsWay H1 coil for 95%. The BrainsWay protocol entailed 18-Hz stimulation at 120% of rMT for 1980 pulses daily for 36 consecutive treatments. A small minority (∼2%) of patients were switched from BrainsWay for tolerability. In these cases, protocols included 1-Hz or continuous TMS on the right, intermittent theta-bust, or bilateral. All Butler patients were outpatient, while 116 of the 630 at McLean began as inpatients.

### Clinical assessment

Clinical response was defined as a decrease in score by ≥50% from baseline to post-treatment. Remission was defined by a post treatment score ≤14 on the IDS-SR and ≤5 on the QIDS. As all QIDS items are included within the IDS-SR, a comparative QIDS score was also calculated for the Butler dataset. The three insomnia-related questions were determined as items 1 (Falling asleep), 2 (Waking up during the night and difficulty falling back to sleep), and 3 (Waking up too early). Each question had a score range from 0-3, with 0 representing no sleep disturbance (ex. “I never take longer than 30 minutes to fall asleep”) and 3 representing the most severe (ex. “I take more than 60 minutes to fall asleep, more than half the time”). The scores of these three questions were summated to create an “insomnia score” with a range from 0-9. Item 4 (Sleeping too much) was not included to separate insomnia from hypersomnia-like sleep disturbances. We then calculated an IDS-SR_25_ total score excluding insomnia items for analyses comparing insomnia to other depressive symptoms rated on the same scale, and likewise for QIDS to create a QIDS_13_.

### Statistical Analysis

Categorical (responders, remitters) and continuous (percent and raw change in baseline to endpoint for IDS-SR, QIDS, and insomnia score) outcomes were explored with descriptive statistics. IDS-SR, QIDS, and insomnia scores were not normally distributed. Wilcoxon Signed Rank tests were used to determine differences in means within sample (baseline to post comparisons) and Mann-Whitney U tests were used to determine differences between sleep quality or medication groups. Statistical significance was defined at p<.05 and was two tailed. Multiple comparisons were Bonferroni corrected. We also analyzed correlations in sleep score with change in overall symptom improvement using Spearman correlation tests.

To determine whether baseline sleep scores predicted TMS outcome, we performed a multilevel logistic regression using the lme4 package (v1.1.33;(Bates D, 2015) with response as categorical outcome variable and baseline sleep, age, sex, and inpatient/outpatient status as fixed effects. Hospital site was included as a random effect to account for a potential influence of variance in the structured data. The above analysis was repeated with remission as outcome variable.

To create a categorial variable “sleep quality,” we chose the median score 4 as the cutoff score based on the histogram and descriptive statistics of baseline insomnia score. Patients with baseline insomnia score with less than or equal to 4 were coded as 0 and those with a score greater than 4 were coded as 1. We then compared IDS-SR_25_/QIDS_18_ scores between no/low insomnia group and high insomnia group.

Furthermore, we examined whether being prescribed sleep medications was associated with improvement in 1) sleep and 2) depression. Patients at both sites were instructed to keep medications stable through TMS. For Butler patients, we filtered through medication lists recorded on the first day of TMS treatment and those taking hypnotics during TMS were coded “1” and those not taking hypnotics were coded “0.” The following generic medications as well as their corresponding brand names were included in our list of sleep-aids: doxepin (Sinequan, Silenor), ramelteon (Rozerem), temazepam (Restoril), triazolam (Halcion), zaleplon (Sonata), zolpidem (Ambien, Zolpimist, Edluar, Intermezzo), eszopiclone (Lunesta), suvorexant (Belsomra). Three antidepressants-trazadone (Desyrel), amitriptyline (Elavil), and mirtazapine (Remeron) were also included, along with melatonin. Anti-anxiety medications were not included in our filter.

To assess whether patients with insomnia taking hypnotic medications had clinical responses to TMS comparable to those not taking hypnotics, we categorized patients into four groups by sleep quality x hypnotic medication use: 1) No/low insomnia and not using hypnotics (“-Insomnia -Meds”) 2) no/low insomnia using sleep meds (“-Insomnia +Meds”) 3) high insomnia and not using hypnotics (“+Insomnia - Meds”) 4) high insomnia despite use of hypnotics (+-Insomnia +Meds”). Kruskal Wallis Test was used to determine differences in baseline and post treatment IDS-SR and insomnia scores between the four groups. To analyze scores across time, mixed repeated measures ANOVA was applied. All statistical analysis was done in R (v4.3.1; R Core Team 2021).

## Results

### Demographics

Between August 2016 and July 2022, 353 patients completed a baseline and post rTMS questionnaire at Butler Hospital, and 630 patients between October 2017 and April 2023 at McLean Hospital. Demographic and clinical characteristics are presented in Table 1. There was a significant decrease in QIDS score from baseline to post-treatment scores across sites (Z=−24.3, p<.001, Fig. 1A).

**Fig 1.**
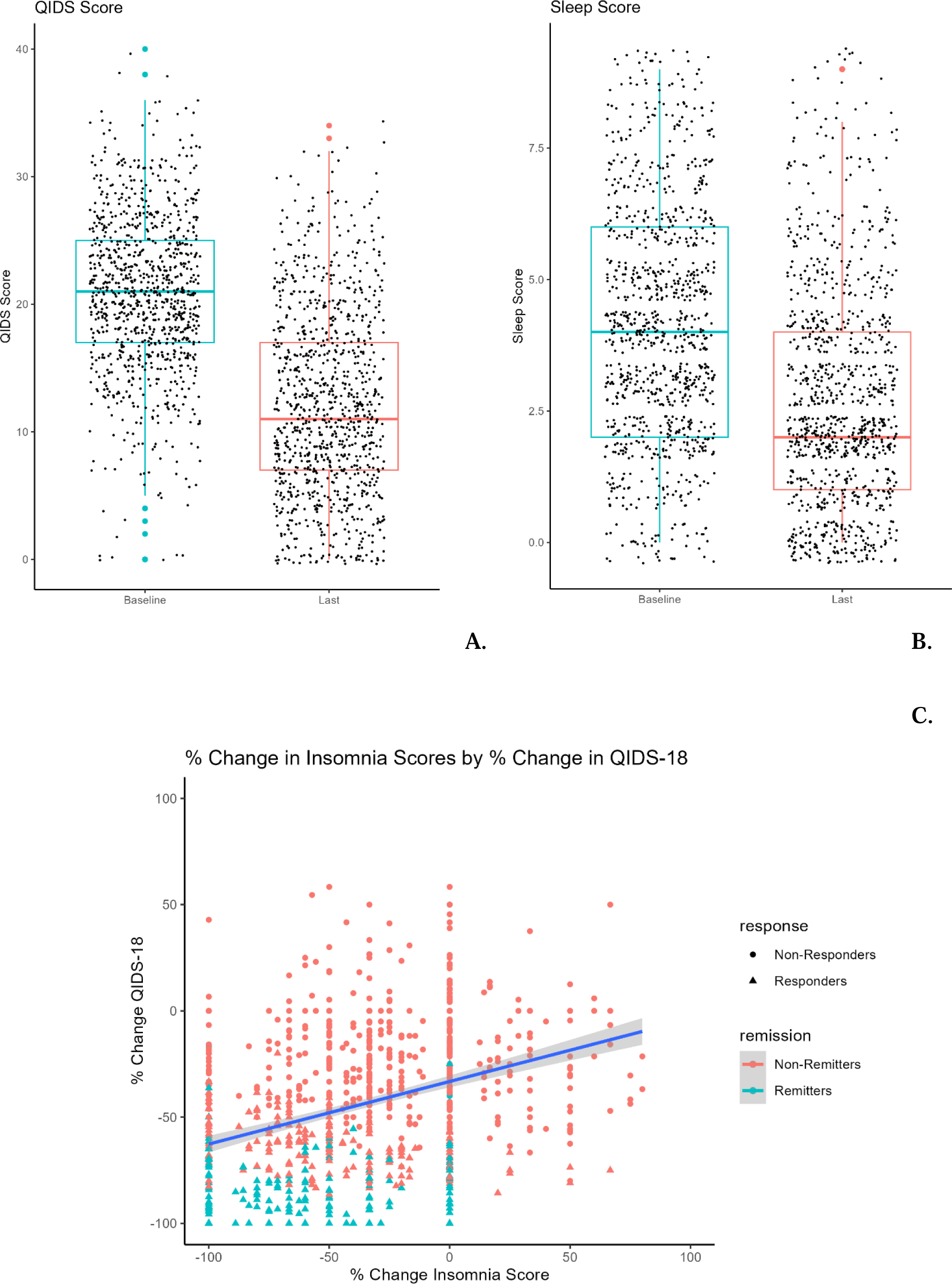
(A) Pre to post-treatment QIDS score with standard deviation (error bars) and individual data points. Pre (20.8±6.18), Post (12.2±7.24). (B) Pre to post-treatment insomnia score with standard deviation (error bars) and individual data points. Pre (4.33±2.10), Post (2.88±2.00). (C) Change in insomnia score positively correlates with change in QIDS_18_ score pre to post TMS.

**Table 1.** Demographic and Clinical Characteristics of three datasets. ^a^ *p* refers to results of Mann-Whitney U test (1) or chi-square test (2) difference between Butler and McLean sample.

| **Variable** | **Combined** | **Butler** | **McLean** |
| --- | --- | --- | --- |
| n | 350 | 350 | 630 |
| Sex (% female) | 63.9% | 69.4% | 60.79% |
| Age (in years) | 48.5 ±17.5 | 46.2±15.6 | 49.8±18.4 |
| Responders (%) | 40.9% | 44.5% | 38.5% |
| Remitters (%) | 21.7% | 25.5% | 19.6% |
| Pre-Tx QIDS Score | 20.8±6.18 | 22.8±5.60 | 19.7±6.21 |
| Post-Tx QIDS Score | 12.2±7.24 | 12.9±7.62 | 11.9±7.00 |
| Percent Change QIDS | -40.7±32.9 | -42.0±30.9 | -38.4±34.0 |
| Pre-Tx Insomnia Score | 4.33±2.10 | 4.60±2.25 | 3.86±2.19 |
| Post-Tx Insomnia Score | 2.88±2.00 | 2.83±2.27 | 2.78±2.01 |

### Effect of TMS on Insomnia

Average baseline insomnia score was 4.33 ± 2.10. We found no significant differences by sex or age. Patients with greater baseline insomnia tended to have greater baseline QIDS scores, i.e., more severe depression (r=.57, *p*<.001). Insomnia scores significantly improved over course of rTMS treatment (Z=−16.45, *p<*.001, Figure 1B).

After responses to the three insomnia-related sleep questions were subtracted, we found percent change in insomnia score correlated in a positive direction with percent change in QIDS_18_ (r=.318, *p*<.001, Fig. 1C), indicating that sleep improved alongside general improvement in depression after TMS.

### Baseline Insomnia and TMS Clinical Outcome

To determine whether baseline insomnia was associated with TMS response or remission rates, we first examined whether baseline and final insomnia/QIDS scores differed between responders and non-responders. Responders had a significantly higher initial insomnia score than non-responders (Z=−2.54, *p*=.011). After TMS, responders had significantly lower insomnia scores than non-responders (Z=−12.64, *p*<.001), as well as greater decrease in insomnia score as measured by percent change (Z=−14.42, p<.001). Likewise, we found a difference in pre-treatment insomnia score between remitters and non-remitters; however, remitters had significantly lower insomnia scores at baseline (Z= −3.11, p=.002) and after TMS (Z=−13.84, p<.001).

A binary logistic multilevel model (MLM) with responder status as dependent variable and baseline insomnia, sex, age, and inpatient status as fixed effects and controlled for location initially showed significant effect of baseline insomnia. However, this effect did not survive upon the addition of baseline QIDS_18_ score. Similarly, a logistic regression model using baseline sleep to predict remission status with the same covariates showed no significant effect of insomnia on remission outcome when QIDS_18_ was included. Baseline insomnia was not a significant predictor of TMS treatment outcome.

### Insomnia as a Modulator of TMS Outcome

We went on to explore whether baseline insomnia influences the trajectory of symptom improvement. Using the Butler data set, insomnia and IDS-SR_25_ scores were plotted every 5 TMS sessions by better and worse sleepers. We found that patients with initially no/minimal insomnia have consistently lower scores for both insomnia and IDS-SR_25_ score across treatment course (Fig. 2A, 2B). This difference was found to be significant by mixed repeated measures ANOVA, which produced significant effects of time (F(3.46, 443)=142.2, p<.001, η^2^= .03), quality (F(1,128)=5.176, p=.025, η^2^) = .23, but not time by quality interaction.

**Figure 2.**
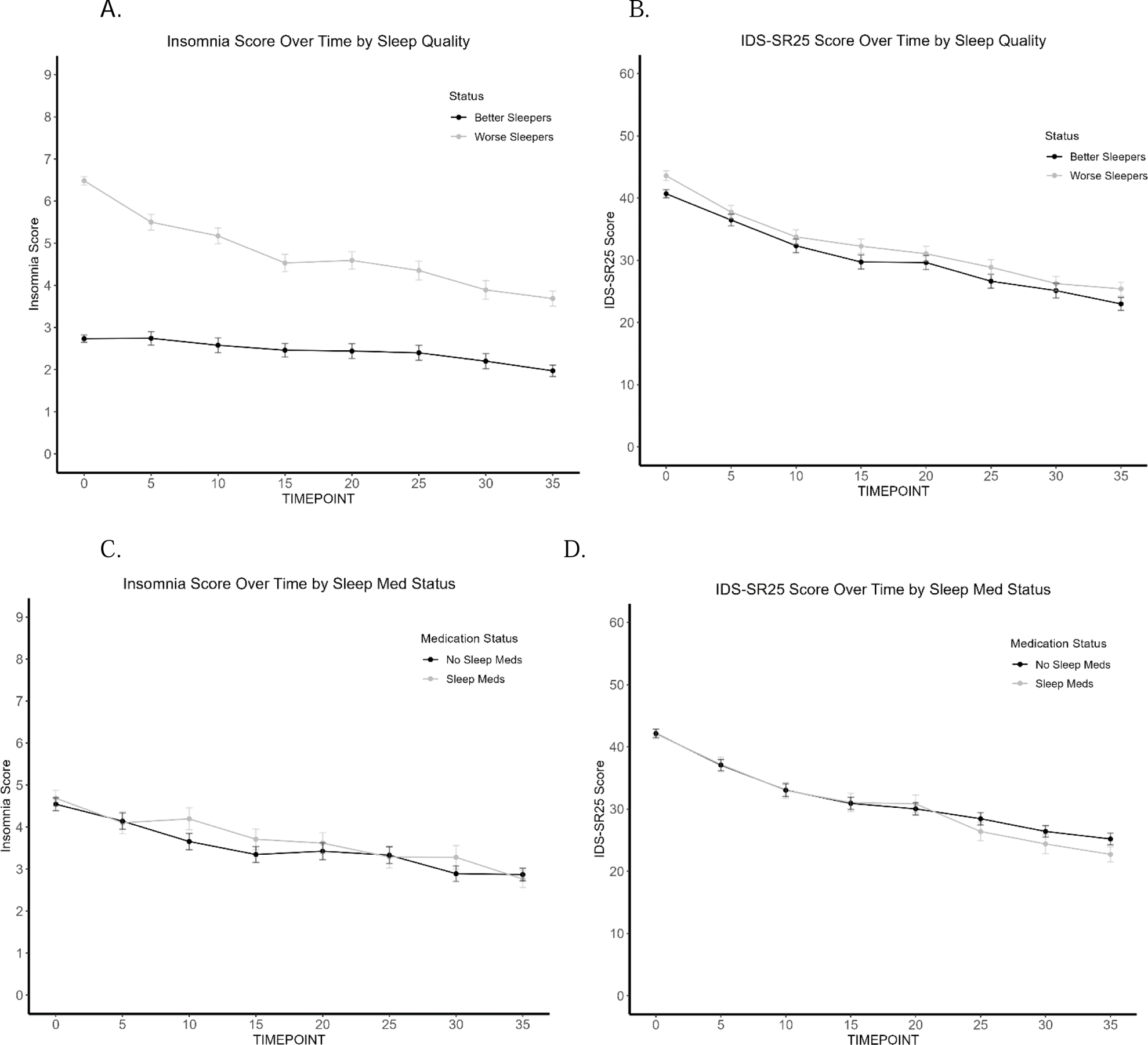
Trajectory of Insomnia and IDS-SR_25_ score over TMS treatment course (Butler). **(A)** Worse sleepers have consistently worse sleep scores across TMS treatment course. **(B)** Worse sleepers have consistently worse IDS-SR_25_ scores across TMS treatment course. **(C)** No difference in insomnia scores across TMS treatment course by sleep medication use. **(D)** No difference in IDS-SR_25_ scores across TMS treatment course by sleep medication use.

### Sleep Medications

To account for the use of hypnotics and their impact on sleep in our naturalistic sample, we found that patients taking sleep medications did not have significantly different baseline/final insomnia or IDS-SR_25_ scores when compared to patients not taking sleep medications. Fig. 2C and 2D show no separation in sleep scores and overall depression scores respectively for the two sleep groups Mixed repeated measures ANOVA revealed a significant effect of only time for dependent measure IDS-SR score (F(3.53, 451.46)=140.82, p<.001, η^2^)= .22) and insomnia score (F(4.76, 609.44)=20.02, p<.001, η^2^)=.04). Regardless of whether hypnotics were used, TMS treatment improved insomnia and other depression symptoms. Sleep medication status was not associated with response or remission outcome, based on chi-square analysis.

### Insomnia and Medication Use

Finally, we sought to determine whether hypnotics influence treatment outcome through modulation of sleep. Table 2 shows the number of patients in each group, mean sleep score, and mean IDS-SR_25_ score before and after TMS. Mixed ANOVA reveals that within patients with no/minimal insomnia, there is no difference in insomnia scores between patients taking sleep medications and patients that are not (F(1, 76)= .208, *p*=.65). Similarly, we find no difference in scores within patients with insomnia by medication status (F(1,72)= .115, *p*=.736).

**Table 2.** Breakdown of the Four Insomnia by Sleep Medication Use Groups. Number per group, and mean scores for each measure. BL= Baseline; IDS-SR25= Inventory of Depressive Symptomology (Self-Report) without sleep items; “-Insomnia -Meds” = No/minimal insomnia without hypnotics; “-Insomnia +Meds” = No/minimal insomnia with hypnotics; “+Insomnia -Meds”= Insomnia without hypnotics; “+Insomnia -Meds”= Insomnia with hypnotics”

| Status | N | BL Insomnia Score | Last Insomnia Score | BL IDS-SR25 Score | Last IDS-SR25 Score |
| --- | --- | --- | --- | --- | --- |
| -Insomnia -Meds | 144 | 2.82 | 2.10 | 41.0 | 23.2 |
| -Insomnia +Meds | 63 | 2.59 | 1.73 | 40.1 | 22.6 |
| +Insomnia -Meds | 95 | 6.62 | 3.80 | 43.6 | 27.6 |
| +Insomnia +Meds | 81 | 6.32 | 3.56 | 43.6 | 22.8 |

Examining IDS-SR_25_ scores over time by sleep quality group (no/low vs. high insomnia) and hypnotic medication use status (Fig. 3A) revealed that once again patients taking hypnotics demonstrated no significant difference to those with healthy sleep and not taking hypnotics. Even more interestingly, while the high insomnia group taking hypnotics had significantly higher baseline IDS-SR scores than patients with no/low initial insomnia, all three groups ended at comparable end depression scores. This is despite demonstrating consistently greater insomnia scores, even by the end of treatment. In contrast, patients with high insomnia not taking medications had significantly higher IDS-SR_25_ scores post-TMS than the other three groups (Fig. 3B). We found an overall significant effect of group (F(3,4)=7.37, p= .042, η^2^)=.79, time (F(7,28)=3.28, p=.011, η^2^)=.21, but not group x time interaction, with significant Kruskal-Wallis at final time point 35 (*p*=.04). Patients using sleep medications appear to have greater improvement in overall depression symptoms, even if insomnia does not significantly improve.

**Figure 3.**
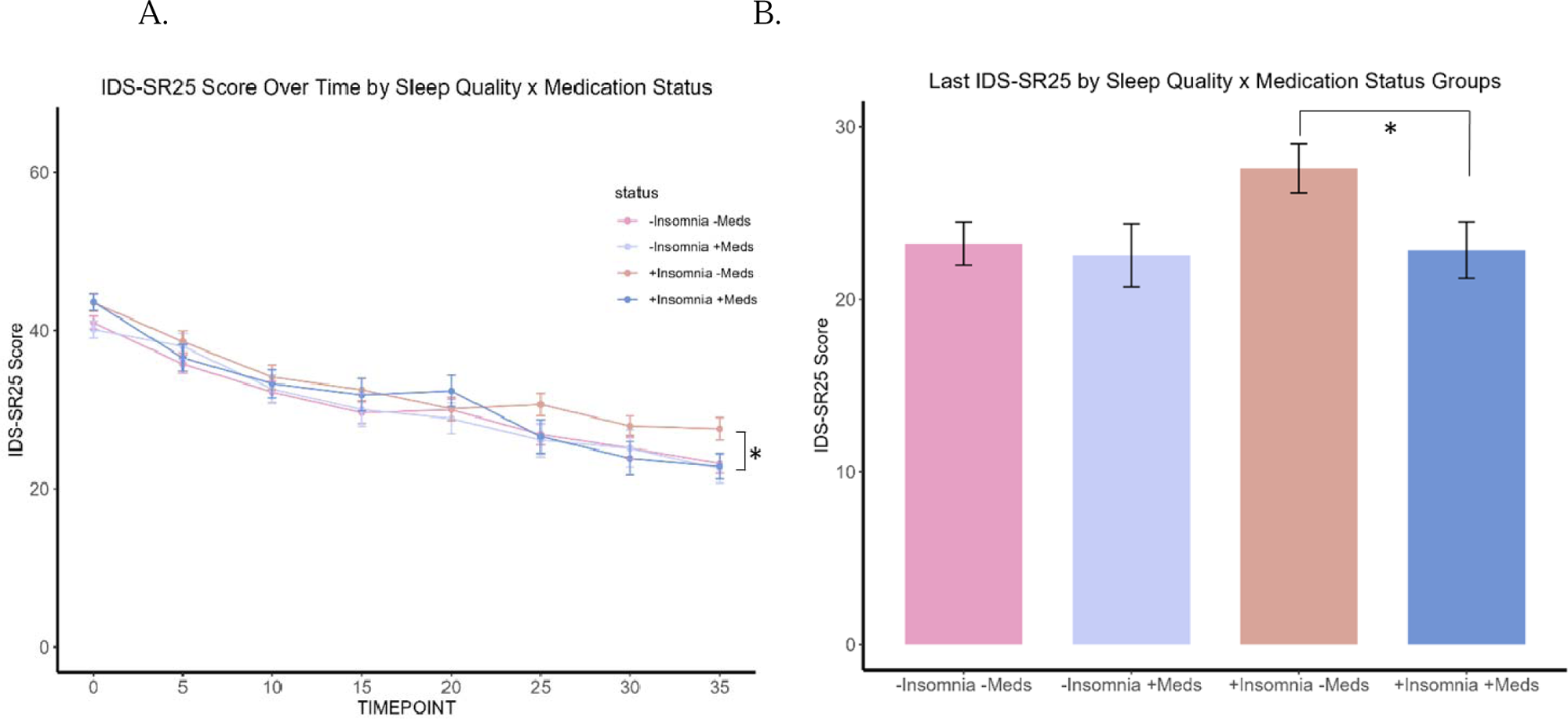
Trajectory of patients by the four insomnia and medication use groups. **(A)** Significant effect of group (*p*=.04) and time (*p*=.01), but not group x time interaction. **(B)** “+Insomnia -Meds” group significantly higher final IDS-SR25 score at timepoint 35.

## Discussion

We present a large, naturalistic study examining the effect of TMS treatment on self-reported insomnia and vice versa in patients with treatment resistant depression. We found that insomnia measured by three items on the IDS-SR/QIDS significantly improved after TMS and improved alongside non-insomnia depression symptoms. While we found differences in baseline insomnia score by responder and remission status, baseline insomnia was not statistically associated with post-treatment status as a responder or remitter, a result that aligns with the majority of other studies that have examined sleep as a predictor of response and found no relation between the two measures (Brakemeier et al., 2008; Fregni et al., 2006; Lowe et al., 2013).

Examining both insomnia score and IDS-SR_25_ score over the course of TMS every 5 sessions revealed that patients with no/low insomnia continue to have consistently better sleep than those with worse baseline sleep. The former group also demonstrated consistently lower IDS-SR_25_ scores, supporting the notion that greater baseline insomnia score is associated with worse depression. The degree of improvement with TMS does not appear to differ, however, as a function of baseline presence of insomnia. This suggests that sleep improves along with other symptoms comprising the depressive syndrome.

Importantly, treating insomnia with hypnotic medications may benefit overall depression improvement even if subjective sleep measures do not significantly improve. Upon categorizing patients by severity of baseline insomnia symptoms and whether they were using hypnotics, we found that within severity groups there was no difference by sleep medication use. For patients with no/low insomnia using hypnotic medication, results suggest that the drugs successfully control insomnia symptoms to produce sleep ratings no different to patients without sleep. While we found no difference in baseline insomnia scores between those with high insomnia by hypnotic use, we start to see a separation over halfway through TMS treatment where patients using sleep medications show a drop in IDS-SR score and end at a position similar to better sleepers. Worse sleepers not using hypnotics display significantly worse final depression scores than all three other groups, suggesting that while sleep issues may persist despite medication use, treating sleep appears to benefit the degree of improvement in depression from TMS. Therefore, it does not appear that patients with poor sleep will always do worse than their better sleeping counterparts; modulating sleep with appropriate medications alongside TMS may improve depression, even if self-reported sleep itself does not significantly improve.

One potential underlying link between sleep, depression, and TMS treatment is plasticity. Impairments in the brain’s ability to reorganize and respond to changing stimuli has been implicated in MDD, such as synaptic depression in the prefrontal cortex (Wilkinson et al., 2019). Interestingly, sleep has been found to modulate synaptic plasticity. For example, in a study using high-frequency electrical stimulation in the motor cortex of rats to induce synaptic plasticity, effects were partially occluded after prolonged wakefulness and restored after sleep (Vyazovskiy et al., 2008). Similar results were found in humans; one night of sleep deprivation blunted the facilitatory effect of paired-associative stimulation (PAS) on motor-evoked potentials found after a normal night’s sleep (Kuhn et al., 2016). The same study also found increased theta power in electroencephalography (EEG) during wake the day after sleep deprivation, proposed to reflect “synaptic weight” or the net synaptic strength during wakefulness. These results are in line with the synaptic homeostasis hypothesis, which theorizes that slow wave activity reflects the synaptic changes generated during wakefulness based on the observed reduction of dendritic spines and synaptic markers during sleep (Centorino et al., 2020). It may be possible therefore that patients with depression and more severe sleep disturbances have further impaired plasticity processes that hinder the TMS-related improvement. Following this logic, improving sleep through medications such as hypnotics may in turn restore plasticity mechanisms and facilitate rTMS treatment.

Several meta-analyses have reported that concurrent cognitive-behavioral therapy for insomnia (CBT-I) may improve the efficacy of anti-depressant treatment (Cunningham & Shapiro, 2018; Gebara et al., 2018; Sweetman et al., 2021). No studies have yet examined targeted insomnia treatment during rTMS, with the exception of one open-label feasibility trial with 2 patients undergoing a 36 day 10-Hz (3,000 pulses) with six weekly 1-hour manualized CBT-I sessions (Norred et al., 2021). Both patients experienced significant improvement in subjective sleep rating and reached remission as measured by the 24-item Hamilton Rating Scale for Depression-24 (HRSD_24_). Our findings may provide support that treating sleep alongside TMS treatment may produce additive antidepressant effects. Future work is needed to determine if improving sleep improves TMS outcomes, both with medications and non-pharmaceutical interventions such as cognitive-behavior therapy for insomnia.

Our interpretations are complicated by a number of confounding variables that cannot be controlled with retrospective, naturalistic studies, such as the wide variety of medication regimes that all have different effects on sleep. We also did not include in our filter criteria other drugs that are not strictly categorized as sleep medications but commonly used to treat sleep, such as benzodiazepines or marijuana. As a result, it is difficult to definitively claim that treating sleep with medications improves TMS outcome. Sleep medications also impact motor threshold and may impact the intensity of stimulation delivered for TMS treatment.

A small percentage of the sample received predominantly 1 Hz, 5 Hz rTMS, or iTBS, which may have different effects on sleep and depression. An exploration of the potential relationship between TMS parameters and sleep is necessary. Another limitation is that we used item-level questions within the IDS-SR to measure sleep, as opposed to a separate scale. The creation of the dichotomous “no/low” and “high” insomnia groups using the mean insomnia score of 4, in line with the method used in Fava et al. (2002), may not be an accurate categorization of insomnia severity (Fava et al., 2002). An individual who scored a 3 on a single item would be categorized by our methods as “no/low” insomnia, though endorsing for example, “I take more than 60 minutes to fall asleep, more than half the time.” As insomnia involves different dimensions of sleep, and there are interindividual differences in phenotype, binary categorization of severity presents a challenge. Future studies may consider creating more than two groups to allow for more granular analyses. All measures of sleep were also subjective, and past studies have previously reported discrepancies between subjective and objective ratings of sleep quality. Additional measures, such as actigraphy, EEG recordings, or biomarkers would offer additional support or insight into the biological underpinnings of the relationship between sleep, TMS, and depression.

## Data Availability

All data produced in the present study are available upon request to the authors

## References

1. Andreescu, C., Mulsant, B. H., Houck, P. R., Whyte, E. M., Mazumdar, S., Dombrovski, A. Y., Pollock, B. G., & Reynolds, C. F., 3rd. (2008). Empirically derived decision trees for the treatment of late-life depression. Am J Psychiatry, 165(7), 855–862. 10.1176/appi.ajp.2008.07081340

2. Bates D, M. M., Bolker B, Walker S. (2015). “Fitting Linear Mixed-Effects Models Using lme4.”. Journal of Statistical Software, 67(1), 1–48. doi:10.18637/jss.v067.i01.

3. Brakemeier, E. L., Luborzewski, A., Danker-Hopfe, H., Kathmann, N., & Bajbouj, M. (2007). Positive predictors for antidepressive response to prefrontal repetitive transcranial magnetic stimulation (rTMS). J Psychiatr Res, 41(5), 395–403. 10.1016/j.jpsychires.2006.01.013

4. Brakemeier, E. L., Wilbertz, G., Rodax, S., Danker-Hopfe, H., Zinka, B., Zwanzger, P., Grossheinrich, N., Varkuti, B., Rupprecht, R., Bajbouj, M., & Padberg, F. (2008). Patterns of response to repetitive transcranial magnetic stimulation (rTMS) in major depression: replication study in drug-free patients. J Affect Disord, 108(1-2), 59–70. 10.1016/j.jad.2007.09.007

5. Brown, J. C., Higgins, E. S., & George, M. S. (2021). Synaptic Plasticity 101: The Story of the AMPA Receptor for the Brain Stimulation Practitioner. Neuromodulation. 10.1016/j.neurom.2021.09.003

6. Carney, C. E., Edinger, J. D., Kuchibhatla, M., Lachowski, A. M., Bogouslavsky, O., Krystal, A. D., & Shapiro, C. M. (2017). Cognitive Behavioral Insomnia Therapy for Those With Insomnia and Depression: A Randomized Controlled Clinical Trial. Sleep, 40(4). 10.1093/sleep/zsx019

7. Carpenter, L. L., Janicak, P. G., Aaronson, S. T., Boyadjis, T., Brock, D. G., Cook, I. A., Dunner, D. L., Lanocha, K., Solvason, H. B., & Demitrack, M. A. (2012). Transcranial magnetic stimulation (TMS) for major depression: a multisite, naturalistic, observational study of acute treatment outcomes in clinical practice. Depress Anxiety, 29(7), 587–596. 10.1002/da.21969

8. Caulfield, K. A., & Brown, J. C. (2022). The Problem and Potential of TMS’ Infinite Parameter Space: A Targeted Review and Road Map Forward. Front Psychiatry, 13, 867091. 10.3389/fpsyt.2022.867091

9. Centorino, M. B., Bajor, L. A., Gootam, P. K., Nakase-Richardson, R., & Kozel, F. A. (2020). The Relationship of Transcranial Magnetic Stimulation With Sleep and Plasticity. J Psychiatr Pract, 26(6), 434–443. 10.1097/PRA.0000000000000506

10. Cunningham, J. E. A., & Shapiro, C. M. (2018). Cognitive Behavioural Therapy for Insomnia (CBT-I) to treat depression: A systematic review. J Psychosom Res, 106, 1–12. 10.1016/j.jpsychores.2017.12.012

11. Dew, M. A., Reynolds, C. F., 3rd, Houck, P. R., Hall, M., Buysse, D. J., Frank, E., & Kupfer, D. J. (1997). Temporal profiles of the course of depression during treatment. Predictors of pathways toward recovery in the elderly. Arch Gen Psychiatry, 54(11), 1016–1024. 10.1001/archpsyc.1997.01830230050007

12. Dunner, D. L., Aaronson, S. T., Sackeim, H. A., Janicak, P. G., Carpenter, L. L., Boyadjis, T., Brock, D. G., Bonneh-Barkay, D., Cook, I. A., Lanocha, K., Solvason, H. B., & Demitrack, M. A. (2014). A multisite, naturalistic, observational study of transcranial magnetic stimulation for patients with pharmacoresistant major depressive disorder: durability of benefit over a 1-year follow-up period. J Clin Psychiatry, 75(12), 1394–1401. 10.4088/JCP.13m08977

13. Edinoff, A. N., Hegefeld, T. L., Petersen, M., Patterson, J. C., 2nd, Yossi, C., Slizewski, J., Osumi, A., Cornett, E. M., Kaye, A., Kaye, J. S., Javalkar, V., Viswanath, O., Urits, I., & Kaye, A. D. (2022). Transcranial Magnetic Stimulation for Post-traumatic Stress Disorder. Front Psychiatry, 13, 701348. 10.3389/fpsyt.2022.701348

14. Fang, H., Tu, S., Sheng, J., & Shao, A. (2019). Depression in sleep disturbance: A review on a bidirectional relationship, mechanisms and treatment. J Cell Mol Med, 23(4), 2324–2332. 10.1111/jcmm.14170

15. Fava, M., Hoog, S. L., Judge, R. A., Kopp, J. B., Nilsson, M. E., & Gonzales, J. S. (2002). Acute efficacy of fluoxetine versus sertraline and paroxetine in major depressive disorder including effects of baseline insomnia. J Clin Psychopharmacol, 22(2), 137–147. 10.1097/00004714-200204000-00006

16. Fregni, F., Marcolin, M. A., Myczkowski, M., Amiaz, R., Hasey, G., Rumi, D. O., Rosa, M., Rigonatti, S. P., Camprodon, J., Walpoth, M., Heaslip, J., Grunhaus, L., Hausmann, A., & Pascual-Leone, A. (2006). Predictors of antidepressant response in clinical trials of transcranial magnetic stimulation. Int J Neuropsychopharmacol, 9(6), 641–654. 10.1017/S1461145705006280

17. Gaynes, B. N., Rush, A. J., Trivedi, M. H., Wisniewski, S. R., Spencer, D., & Fava, M. (2008). The STAR*D study: treating depression in the real world. Cleve Clin J Med, 75(1), 57–66. 10.3949/ccjm.75.1.57

18. Gebara, M. A., Siripong, N., DiNapoli, E. A., Maree, R. D., Germain, A., Reynolds, C. F., Kasckow, J. W., Weiss, P. M., & Karp, J. F. (2018). Effect of insomnia treatments on depression: A systematic review and meta-analysis. Depress Anxiety, 35(8), 717–731. 10.1002/da.22776

19. George, M. S., Taylor, J. J., & Short, E. B. (2013). The expanding evidence base for rTMS treatment of depression. Curr Opin Psychiatry, 26(1), 13–18. 10.1097/YCO.0b013e32835ab46d

20. Kuhn, M., Wolf, E., Maier, J. G., Mainberger, F., Feige, B., Schmid, H., Burklin, J., Maywald, S., Mall, V., Jung, N. H., Reis, J., Spiegelhalder, K., Kloppel, S., Sterr, A., Eckert, A., Riemann, D., Normann, C., & Nissen, C. (2016). Sleep recalibrates homeostatic and associative synaptic plasticity in the human cortex. Nat Commun, 7, 12455. 10.1038/ncomms12455

21. Lowe, A., Rajaratnam, S. M., Hoy, K., Taffe, J., & Fitzgerald, P. B. (2013). Can sleep disturbance in depression predict repetitive transcranial magnetic stimulation (rTMS) treatment response? Psychiatry Res, 210(1), 121–126. 10.1016/j.psychres.2013.04.028

22. Murphy, M. J., & Peterson, M. J. (2015). Sleep Disturbances in Depression. Sleep Med Clin, 10(1), 17–23. 10.1016/j.jsmc.2014.11.009

23. Norred, M. A., Haselden, L. C., Sahlem, G. L., Wilkerson, A. K., Short, E. B., McTeague, L. M., & George, M. S. (2021). TMS and CBT-I for comorbid depression and insomnia. Exploring feasibility and tolerability of transcranial magnetic stimulation (TMS) and cognitive behavioral therapy for insomnia (CBT-I) for comorbid major depressive disorder and insomnia during the COVID-19 pandemic. Brain Stimul, 14(6), 1508–1510. 10.1016/j.brs.2021.09.007

24. Sunderajan, P., Gaynes, B. N., Wisniewski, S. R., Miyahara, S., Fava, M., Akingbala, F., DeVeaugh-Geiss, J., Rush, A. J., & Trivedi, M. H. (2010). Insomnia in patients with depression: a STAR*D report. CNS Spectr, 15(6), 394–404. 10.1017/s1092852900029266

25. Sweetman, A., Lack, L., Van Ryswyk, E., Vakulin, A., Reed, R. L., Battersby, M. W., Lovato, N., & Adams, R. J. (2021). Co-occurring depression and insomnia in Australian primary care: recent scientific evidence. Med J Aust, 215(5), 230–236. 10.5694/mja2.51200

26. Troxel, W. M., Kupfer, D. J., Reynolds, C. F., 3rd, Frank, E., Thase, M. E., Miewald, J. M., & Buysse, D. J. (2012). Insomnia and objectively measured sleep disturbances predict treatment outcome in depressed patients treated with psychotherapy or psychotherapy-pharmacotherapy combinations. J Clin Psychiatry, 73(4), 478–485. 10.4088/JCP.11m07184

27. Vyazovskiy, V. V., Cirelli, C., Pfister-Genskow, M., Faraguna, U., & Tononi, G. (2008). Molecular and electrophysiological evidence for net synaptic potentiation in wake and depression in sleep. Nat Neurosci, 11(2), 200–208. 10.1038/nn2035

28. Wilkinson, S. T., Holtzheimer, P. E., Gao, S., Kirwin, D. S., & Price, R. B. (2019). Leveraging Neuroplasticity to Enhance Adaptive Learning: The Potential for Synergistic Somatic-Behavioral Treatment Combinations to Improve Clinical Outcomes in Depression. Biol Psychiatry, 85(6), 454–465. 10.1016/j.biopsych.2018.09.004

29. Ziemann, U. (2004). TMS induced plasticity in human cortex. Rev Neurosci, 15(4), 253–266. 10.1515/revneuro.2004.15.4.253

